# Extended-spectrum beta-lactamase-producing *Escherichia coli* and *Klebsiella pneumoniae* from human carriage, the human-polluted environment, and food: molecular epidemiology of two prospective cohorts in five European metropolitan areas

**DOI:** 10.1101/2024.11.21.24316745

**Authors:** TD Verschuuren, J Guther, ME Riccio, D Martak, E Salamanca, S Göpel, N Conzelmann, J Scharringa, P Musicha, IB Autenrieth, BS Cooper, D Hocquet, E Tacconelli, J Rodríguez-Baño, S Harbarth, AC Fluit, S Peter, JAJW Kluytmans

**Affiliations:** Julius Centre for Health Sciences and Primary Care, University Medical Centre Utrecht, Utrecht, the Netherlands; Mahidol-Oxford Tropical Medicine Research Unit, Faculty of Tropical Medicine, Mahidol University, Bangkok, Thailand; Institute of Medical Microbiology and Hygiene, University Hospital Tübingen, Tübingen, Germany; Infection Control Programme and Division of Infectious Diseases, University of Geneva Hospitals and Faculty of Medicine, Geneva, Switzerland; Infection Control Unit, Centre de Ressources Biologiques - Filière Microbiologique de Besançon, Centre Hospitalier Universitaire, Besançon, France; Chrono-environnement UMR 6249, CNRS, Université Bourgogne Franche-Comté, Besançon, France; Department of Infectious Diseases and Clinical Microbiology, University Hospital Virgen Macarena; Department of Medicine, University of Sevilla / Biomedicines Institute of Sevilla (IBiS), Sevilla, Spain; Centro de Investigación Biomédica en Red en Enfermedades Infecciosas (CIBERINFEC), Madrid, Spain; Infectious Diseases, Department of Internal Medicine, University Hospital Tübingen, Tübingen, Germany; Department of Medical Microbiology, University Medical Centre Utrecht, Utrecht, the Netherlands; Centre for Tropical Medicine & Global Health, Nuffield Department of Medicine, University of Oxford, United Kingdom; University Hospital Heidelberg, Heidelberg, Germany; Infectious Diseases, Department of Diagnostics and Public Health, University of Verona, Italy

**Keywords:** multidrug resistance, *Escherichia coli*, *Klebsiella pneumoniae*, genomics, human carriage, human-polluted environment, food

## Abstract

**Objectives:** For 475 ESBL-producing *Escherichia coli* (ESBL-Ec), and 171 ESBL-producing *Klebsiella pneumoniae* (ESBL-Kp) collected from human carriers, the human-polluted (hp)-environment, and food: (i) to compare the antimicrobial resistance gene (ARG) content, and (ii) to assess clonal relationships between human and non-human isolates.

**Methods:** Two prospective multicentre cohorts were assessed: colonised hospitalised index-subjects and household contacts, and long-term care facility (LTCF) residents. Additionally, linked hp-environment and food samples were collected. Presence of ARGs were assessed using pairwise comparisons and proportional similarity index (PSI). Clonal relationships were assessed using cgMLST distance visualisations and maximum likelihood phylogeny.

**Results:** ESBL-Ec and ESBL-Kp co-occurred in 14/65 households, 3/6 LTCFs, and in 33/202 of ESBL-positive participants. Thirty-nine percent of detected ARG types were found in both species (36/93). Frequencies of beta-lactamase, ESBL, aminoglycoside, and sulfonamide ARG types from human ESBL-Ec and ESBL-Kp overlapped considerably: PSIs 0.59-0.75, and were equal or higher compared to the overlap between ESBL-Ec from humans and food isolates: PSIs 0.33-0.72. Isolates from humans and the hp-environment were frequently clonally related, indicating human contamination of the environment. Links with food isolates were observed less frequently. For ESBL-Ec both interregional and regional clonal dissemination were observed, while for ESBL-Kp clonal dissemination was mainly regional.

**Conclusions:** ESBL-Ec and ESBL-Kp from human carriage showed considerable overlap in ARG content. Furthermore, clonal links were observed frequently between humans and hp-environment, and with lower frequency between humans and food. These findings are consistent with human-to-human transmission as an important driver of ARG spread in humans.

## INTRODUCTION

Antimicrobial resistance was declared as one of the top 10 public health threats facing humanity by the World Health Organization in 2019 [1]. In Europe, extended-spectrum beta-lactamase-producing *Escherichia coli* (ESBL-Ec) and *Klebsiella pneumoniae* (ESBL-Kp) are of particular concern, as these species are the most frequent causative agents of infections with antimicrobial resistant pathogens [2]. Furthermore, previous studies have shown that intestinal carriage of ESBL-producing Enterobacterales will result in infection in 8% of carriers [3]. Scientific effort has been put into elucidating reservoirs of human carriage for these bacteria, but the data on the relevance of sources outside humans are conflicting [4–7]. Most antimicrobial resistance genes (ARGs) that have been observed in *E. coli* also occur in *K. pneumoniae*, and vice versa, which is indicative of horizontal gene transfer (HGT) [4, 6, 8, 9]. However, ARG types have rarely been compared systematically between multidrug-resistant *E. coli* and *K. pneumoniae* [10–14]. Five studies have been performed in a retrospective way and were mostly single-centre (*n*=3), with relatively small sample sizes (*n*=71-150) [10–14]. Here, we statistically compare the ARG types occurring in ESBL-Ec and ESBL-Kp from human carriers with a close relationship to healthcare, the human-polluted (hp)-environment, and food. Additionally, we assess clonal relationships between human and non-human isolates.

## METHODS

### Study design, and data-collection

Two prospective cohort studies were performed (2017-19) in five European metropolitan areas: Besançon (France), Geneva (Switzerland), Seville (Spain), Tübingen (Germany), and Utrecht (the Netherlands). Data collection, sample processing, and microbiological methods have been described previously [7, 15]. Briefly, the household study recruited ESBL-Ec- and ESBL-Kp-positive index patients during hospitalisation. Participants were followed up at home for four months after discharge, together with ≥1 other household member, and provided a faecal sample at four time points during follow-up. In the long-term care facility (LTCF) study, participating residents were followed for eight months, and provided a faecal sample or perianal swab at eight time points during follow-up (S1: Figure S1). Additionally, food and hp-environment samples were collected: (i) food was sampled at three time points in supermarkets where participating households reported shopping, and eight times in the kitchen of the participating LTCFs, (ii) samples from the hp-environment were collected from: a) LTCF U-bends, b) LTCF surfaces, c) LTCF wastewater, d) wastewater treatment plant (WWTP) inflow connected to the LTCF, and e) river downstream to the WWTP. Reservoirs a) and b) were sampled twice, the other reservoirs eight times (S1: Figure S1).

### Ethics

Approval was obtained by the ethical review board of each institution. All enrolled participants or their representatives provided written informed consent for participation in this study.

### Microbiological methods

The microbiology laboratory of each participating centre used selective culture media, and selective enrichment broth with standardised methods for the collected human, environmental surface, wastewater, and food samples. Subsequently, identification of ESBL-Ec and ESBL-Kp was performed (S1: Appendix 2).

### Sequencing and selection of unique isolates

Isolates identified as ESBL-Ec or ESBL-Kp were shipped to Tübingen or Utrecht. DNA isolation (DNeasy UltraClean Microbial Kit, Qiagen), sequencing (NextSeq and MiSeq platforms, Illumina, San Diego, USA), and *de novo* assembly (Spades v3.11.1) was performed on all isolates. Quality of assembled sequences was assessed (S1: Appendix 3). In this study, only unique isolates were included (S1: Appendix 3). A list of all included strains, labelled with a public identifier, is provided in the supplement (S2). Original sample IDs are not known by anyone outside the research group.

### *In silico* molecular typing

ARGs conferring resistance to clinically relevant antimicrobials were identified using ResFinder (v3.2) (S1: Appendix 4, Figure S2) [16]. These included ARGs conferring resistance to: (i) penicillins only (small- or broad-spectrum), from here on referred to as beta-lactams, (ii) penicillins and cephalosporins, from here referred to as ESBLs, (iii) penicillins and carbapenems, referred to as carbapenemases, (iv) fluoroquinolones, (v) aminoglycosides, (vi) fosfomycin, (vii) trimethoprim, (viii) sulfonamides, and lastly (ix) colistin. Classifications were made according to genotype-phenotype (antimicrobial and class) translations available in the ResFinder database (https://bitbucket.org/genomicepidemiology/resfinder_db/src/master/phenotypes.txt) and EUCAST Clinical Breakpoint Tables (v13.0) to further subgroup the ‘β-lactam’ class provided by ResFinder [17, 18]. A list of all detected ARG types with classifications is included in the supplement (S1: Appendix 5). Sequence types (STs) were identified using published tools, and sub-clades were assigned to ST131 (ESBL-Ec) isolates (S1: Appendix 4) [19, 20].

### Analysis

The proportions of the 10 most frequently occurring ARG types per class were compared using a two-proportion z-test. The pairwise overlap between sampled reservoirs of acquired ARG type distributions was quantified with Czekanowski’s proportional similarity index [4] *PSI* = 1 − 0. 5 ∑ *k|p*(*reservoir*[*n*])*k* − *q*(*reservoir*[*nx*])*k*|, where *p* corresponded to the relative frequency of gene type *k* in reservoir *n*, and *q* corresponded to the relative frequency of the same gene type in reservoir *nx* [4, 21]. The denominator of the relative frequency was the total number of genes in the corresponding ARG class. The PSI is a proportion, with 0 being interpreted as no overlap, and 1 as perfect overlap in ARG type distributions between two reservoirs. Bootstrap iterations (5,000) were used to calculate 95% confidence intervals (CIs) (boot R-package, v1.3.23) [4]. Clonal transmission between humans was described previously [15]. Here, we assessed clonal relationships between human and non-human isolates, with a visualisation where isolates were ordered based on epidemiological setting (qgraph R-package, v1.9.4). To prevent detection of spurious relationships between human and non-human isolates, a cgMLST threshold was chosen for ESBL-Ec of ≤0.0040 (∼10 alleles) for definition of clonally related pairs, based on previous studies [7, 21]. The threshold for ESBL-Kp was set at ≤0.0035 (∼10 alleles) [21, 22]. A sensitivity analysis with increased thresholds was performed. Lastly, maximum likelihood (ML) trees were created for ESBL-Ec and ESBL-Kp (S1: Appendix 6).

## RESULTS

In total, 196 participants were included (110 household members, and 86 LTCF residents), carrying 205 ESBL-Ec, and 101 ESBL-Kp isolates (Table 1). Furthermore, 232 hp-environment and 108 food isolates were included, consisting of 270 ESBL-Ec, and 70 ESBL-Kp isolates (Table 1). During follow-up, both species were detected in human samples from 14/65 (22%) households, and 3/6 LTCFs, and 17% of participants were colonised with both ESBL-Ec and ESBL-Kp. A mean of 1.6 isolates (range: 1-10) were detected in participants during follow-up (Table 1). On average, participants from Utrecht were colonised with 1.1 unique isolates, while participants from Seville were colonised with 1.7 unique isolates (p<0.0001), mostly due to more frequent carriage of ESBL-Kp isolates.

**Table 1.** Characteristics of the included first unique ESBL-producing *E. coli* and *K. pneumoniae* isolates from humans, the human-polluted environment, and food.

|  | ESBL+ | ESBL-Ec | ESBL-Kp |
| --- | --- | --- | --- |
| **ESBL-positive participants<sup>a</sup> (n)** | 196 | 151 | 78 |
| household members | 110 | 82 | 41 |
| LTCF-residents | 86 | 69 | 37 |
| **Number of unique isolates per participant (mean; range)** | 1.6 (1-10) | 1.1 (0-7) | 0.52 (0-3) |
| household members<sup>b</sup> | 1.4 (1-10) | 1.0 (0-7) | 0.43 (0-3) |
| LTCF residents<sup>c</sup> | 1.7 (1-5) | 1.1 (0-5) | 0.60 (0-3) |
| **Unique isolates (n)** | 646 | 475 | 171 |
| human | 306 | 205 | 101 |
| human-polluted environment | 232 | 175 | 57 |
| food | 108 | 95 | 13 |
| **Composition human-polluted environment (n)** |  |  |  |
| LTCF surface | 15 | 7 | 8 |
| LTCF U-bend | 26 | 13 | 13 |
| LTCF wastewater outflow | 34 | 19 | 15<sup>d</sup> |
| wastewater treatment plant inflow | 73 | 64 | 9 |
| downstream river | 84 | 72 | 12 |

<sup>a</sup>n: number of participants carrying ESBL-*E. coli* or ESBL-*K. pneumoniae* on at least one time-point. <sup>b</sup>The maximum follow-up of a household members was four months. <sup>c</sup>The maximum follow-up of LTCF residents was eight months. <sup>d</sup>seven isolates were retrospectively identified as *K. variicola*.

### Comparison of ARG content

Of a total of 93 detected ARG types, 39% (*n*=36) were detected in both species, demonstrating overlap of resistance genes from all classes, except carbapenems and colistin (Table 2). ARGs encoding for carbapenemases (ESBL-Ec (*n*=2): *bla*_OXA-48_, *bla*_OXA-181_, ESBL-Kp: *bla*_KPC-2_ (*n*=2)) and colistin-resistance (ESBL-Kp: *mcr-1* (*n*=1)) were rare. When assessing the 10 most frequently occurring ARG types per class, we observed similar frequencies for ESBL-Ec and ESBL-Kp from human isolates. However, overall proportions of ARGs were higher for ESBL-Kp (Figure 1 and S1: Figure S3). For example, compared to ESBL-Ec, ESBL-Kp were more likely to harbour ARGs from the *bla*_SHV_ family (77% vs 12%), resistance genes *aac(6’)-Ib-cr* (51% vs 14%), *aac(3)-IIa* (45% vs 8%), and *bla*_OXA-1_ (50% vs 15%), and *fosA* genes (75% vs 1%) (Figure 1 and S1: Figure S3). When assessing the sampled reservoirs separately, ESBL-Ec from food showed differences in detected ARG types compared to other groups. For example, ESBL-Ec from food harboured less *bla*_OXA-1_ (2% vs 28%), *bla*_CTX-M-15_ (12% vs 59%)_,_ *aac(6’)-Ib-cr* (2% vs 27%)*, aac(3)-IIa* (2% vs 21%), and more *bla*_SHV-12_ (39% vs 6%), *bla*_CTX-M-1_ (32% vs 5%), *ant(3”)-Ia* (38% vs 9%), and *sul3* (26% vs 3%) compared to the cumulative frequencies in other groups (Figure 1 and S1: Figure S3).

**Figure 1.**
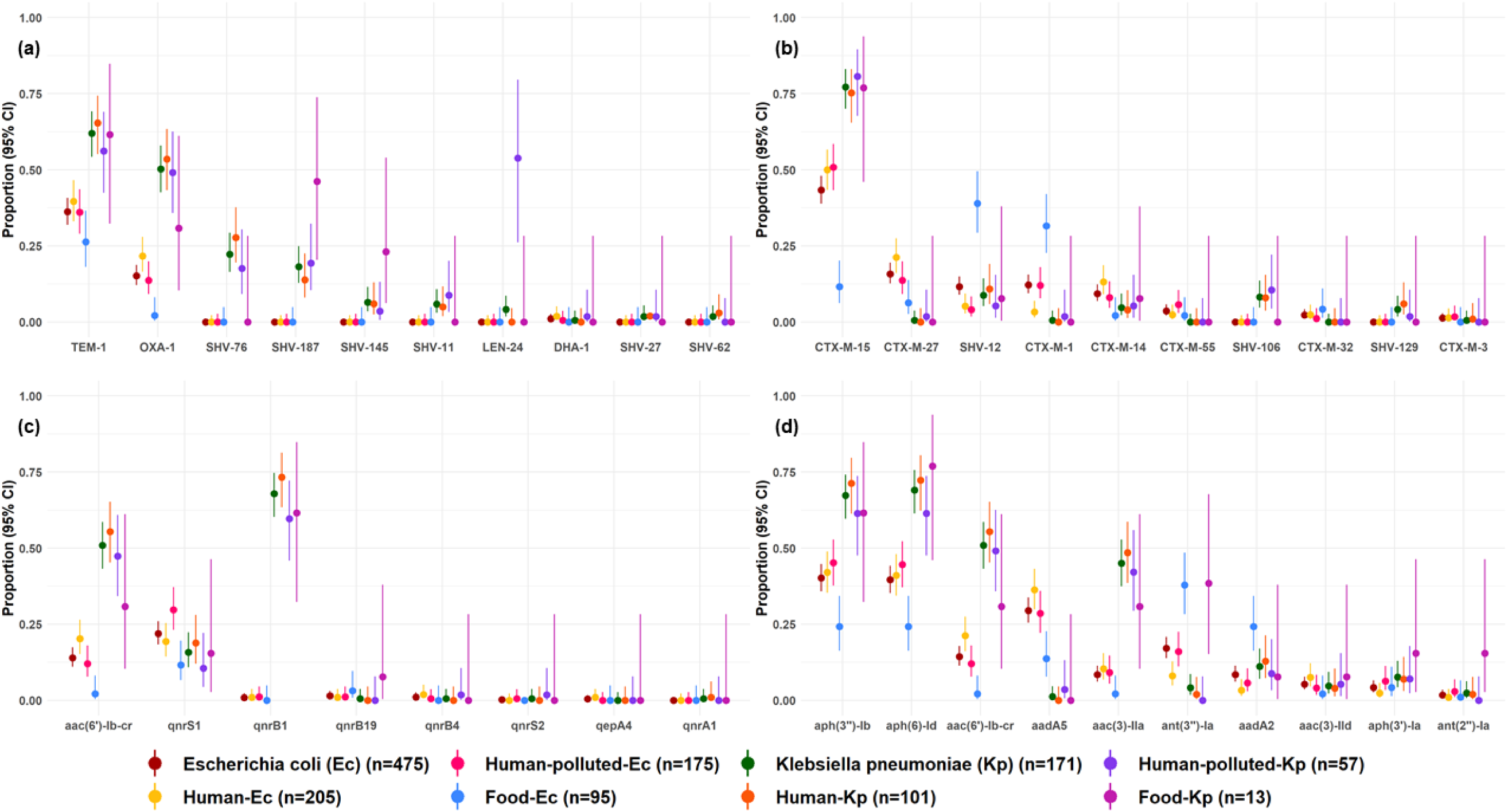
Proportions (95% CI) of the ten most frequent antimicrobial resistance gene (ARG) types per isolate. **Panel A)** beta-lactamases, **Panel B)** ESBLs, **Panel C)** fluoroquinolones, **Panel D)** aminoglycosides. Proportions of the ten most frequent ARG types for fosfomycin, trimethoprim, and sulfonamides are described in the supplement (S1: Fig S3).

**Table 2.**
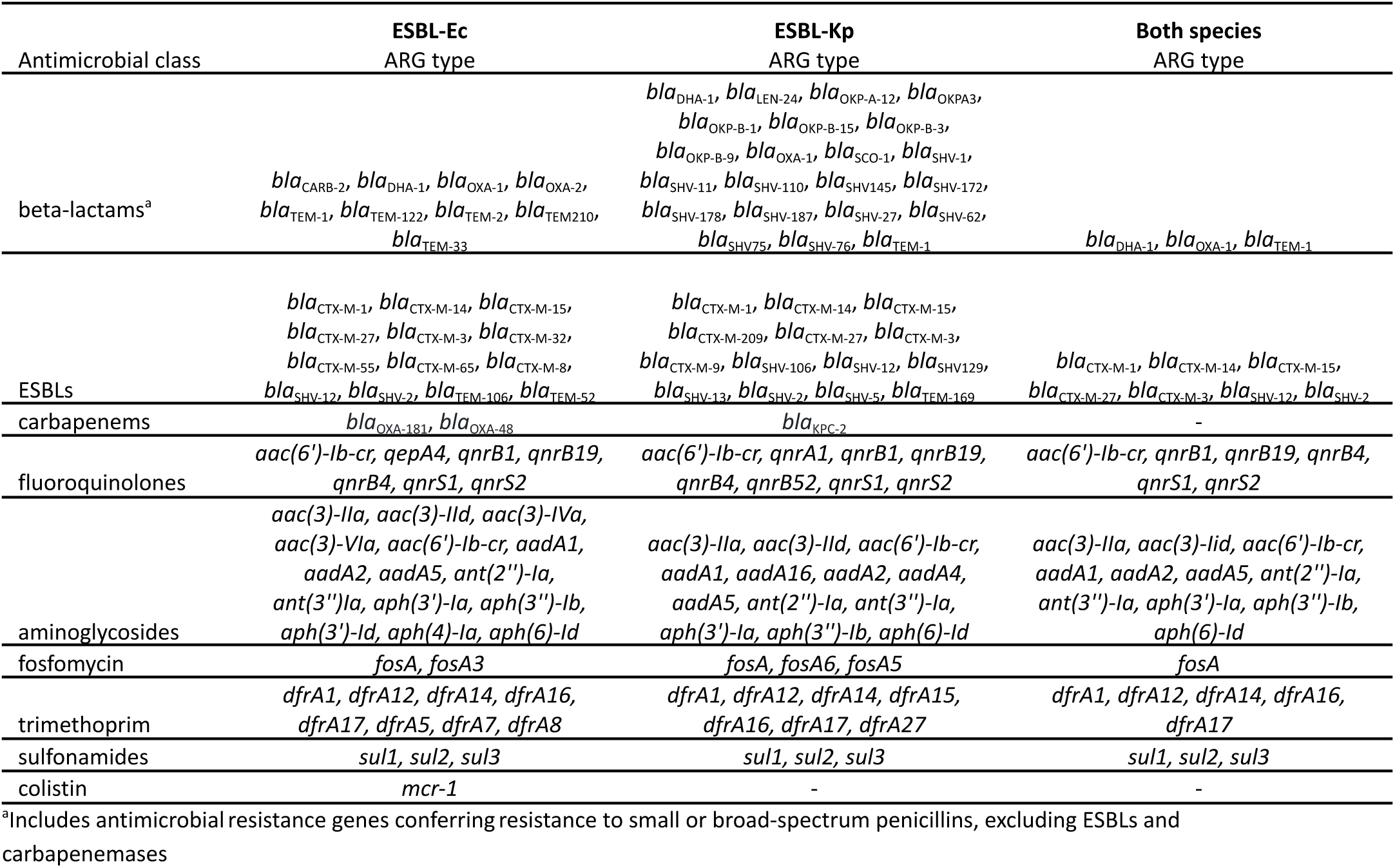
Detected antimicrobial resistance gene (ARG) types grouped per antimicrobial class of conferred resistance.

The PSI was calculated in order to quantify the total overlap of observed ARG types (grouped per antimicrobial class) between the sampled reservoirs. Overall, the ARG types from the same species observed in human and human-polluted environment isolates were most similar, ranging between 0.76 (95% CI 0.7-0.8) for beta-lactams from ESBL-Kp to 0.94 (0.9-1.0) for fosfomycin from ESBL-Ec (Figure 2 and S1: Figure S4). However, ESBL-Ec and ESBL-Kp from humans also showed considerable similarity in beta-lactams (PSI 0.63; 0.5-0.7), ESBLs (PSI 0.59; 0.5-0.7), aminoglycosides (PSI 0.68; 0.6-0.7), and sulfonamides (PSI 0.75; 0.6-0.9) ARG types. In fact, for these ARGs, PSIs were equal or higher to PSIs from humans and food harbouring ESBL-Ec. Lastly, less overlap was observed between ESBL-Ec and ESBL-Kp from humans for fluoroquinolone (PSI 0.37; 0.3-0.5), fosfomycin (PSI 0.24; 0.1-0.3), and trimethoprim (PSI 0.36; 0.3-0.5) ARG types (Figure 2 and S1: Figure S4).

**Figure 2.**
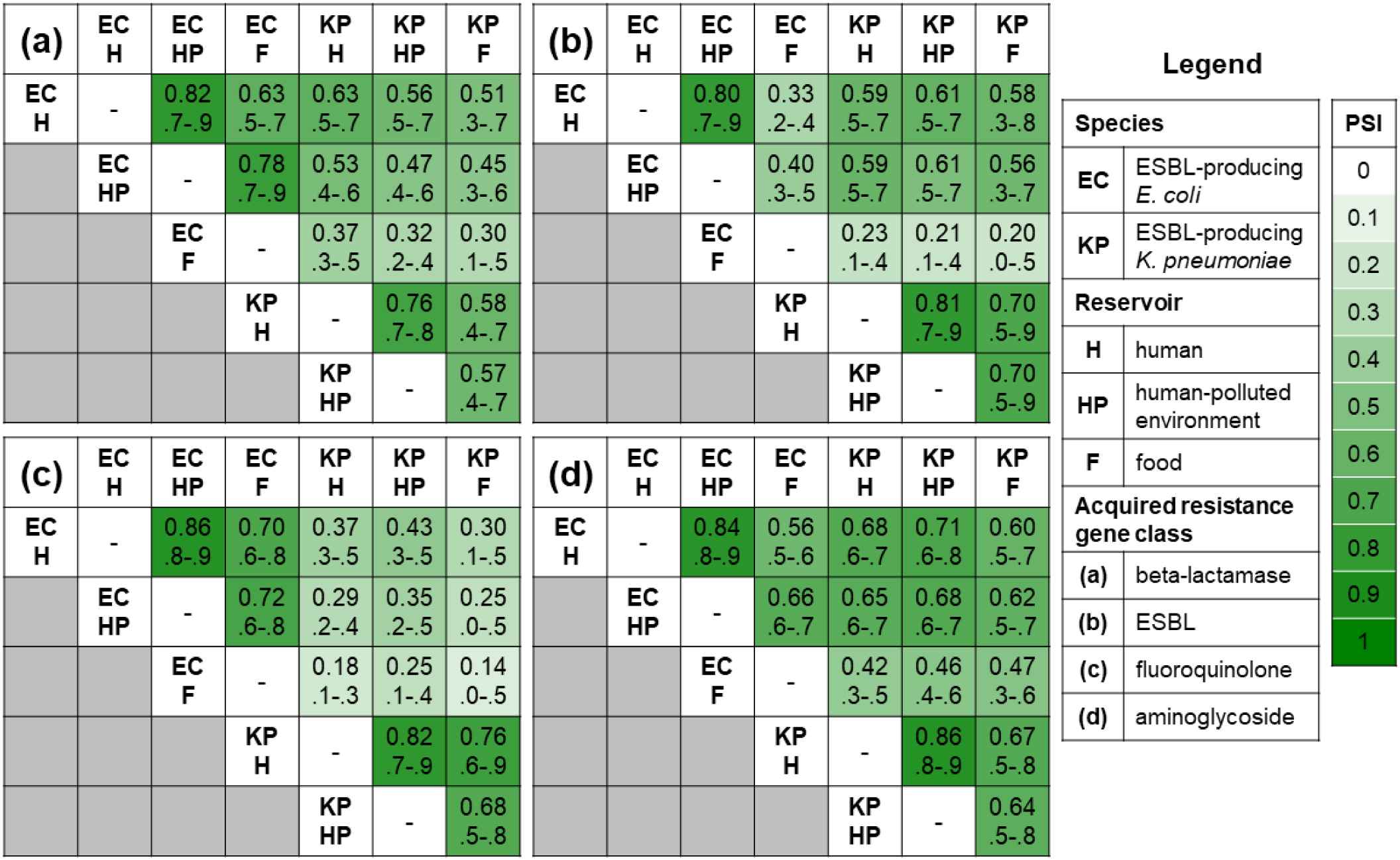
Czekanowski’s proportional similarity index (PSI) (95% confidence interval) of detected antimicrobial resistance gene (ARG) types. **Panel A)** beta-lactamases conferring resistance to penicillins only (small and broad spectrum), **Panel B)** ESBL, **Panel C)** fluoroquinolones, and **Panel D)** aminoglycosides. The PSI is calculated with the following formula: 1 − 0. 5 ∑ *k*|*p*(*reservoir*[*n*])*k* − *q*(*reservoir*[*nx*])*k*|, where *p* corresponded to the relative frequency of gene type *k* (e.g. *bla*_CTX-M-15_) in reservoir *n* (e.g. ESBL-*E. coli* from humans), and *q* corresponded to the relative frequency of the same gene type in reservoir *nx* (e.g. ESBL-*K. pneumoniae* from humans). The numerator of the relative frequency was the count of each ARG type. The denominator of the relative frequency was the total number genes of the corresponding ARG class. The PSI is a proportion, with 0 interpreted as no overlap, and 1 as perfect overlap in ARG type distributions between two reservoirs. 95% confidence intervals were calculated with 5,000 bootstrap iterations. PSI analysis for fosfomycin, trimethoprim, and sulfonamides ARGs are described in the supplement (S1: Fig S4).

### Comparison of core genome content of isolates from humans, the hp-environment, and food

The three most frequent STs within ESBL-Ec were: ST131 (28%; 44% in human isolates, 23% hp-environment, 0% food), ST10 (11%; 10% human, 10% hp-environment, 12% food), ST69 (5.1%; 2.8% human, 5.1% hp-environment, 9.5% food). For ESBL-Kp, the three most frequent STs were ST405 (22%; 27% in human isolates, 18% hp-environment, 0% food), ST307 (10%; 10% human, 12% hp-environment, 0% food), ST323 (5.8%; 3.0% human, 12% hp-environment, 0% food). During follow-up, eight participants were colonised with >1 ST131 isolate, often harbouring different ESBL genes (S1: Table S1).

A pattern of clonally related isolates between humans and the hp-environment was observed (Figure 3a-b). This observation was most pronounced in Seville for both ESBL-Ec and ESBL-Kp, where isolates from residents were regularly related to LTCF surfaces, U-bends, and LTCF wastewater outflow (Figure 3c-d). These connections reflected spread from LTCF residents to the LTCF-associated environments, and potentially human acquisition through contaminated surfaces (and U-bends). Furthermore, similar patterns were observed for ESBL-Ec isolates from Besançon, Tübingen, and Geneva (Figure 3a, and S1: Figure S5). Observed clonally related isolates from humans and WWTP inflow or downstream rivers likely reflected spread of certain genetically conserved strains in a larger geographical area. Lastly, we found two clonally related human-food pairs for ESBL-Ec, both without a known epidemiological link (Figure 3a).

**Figure 3.**
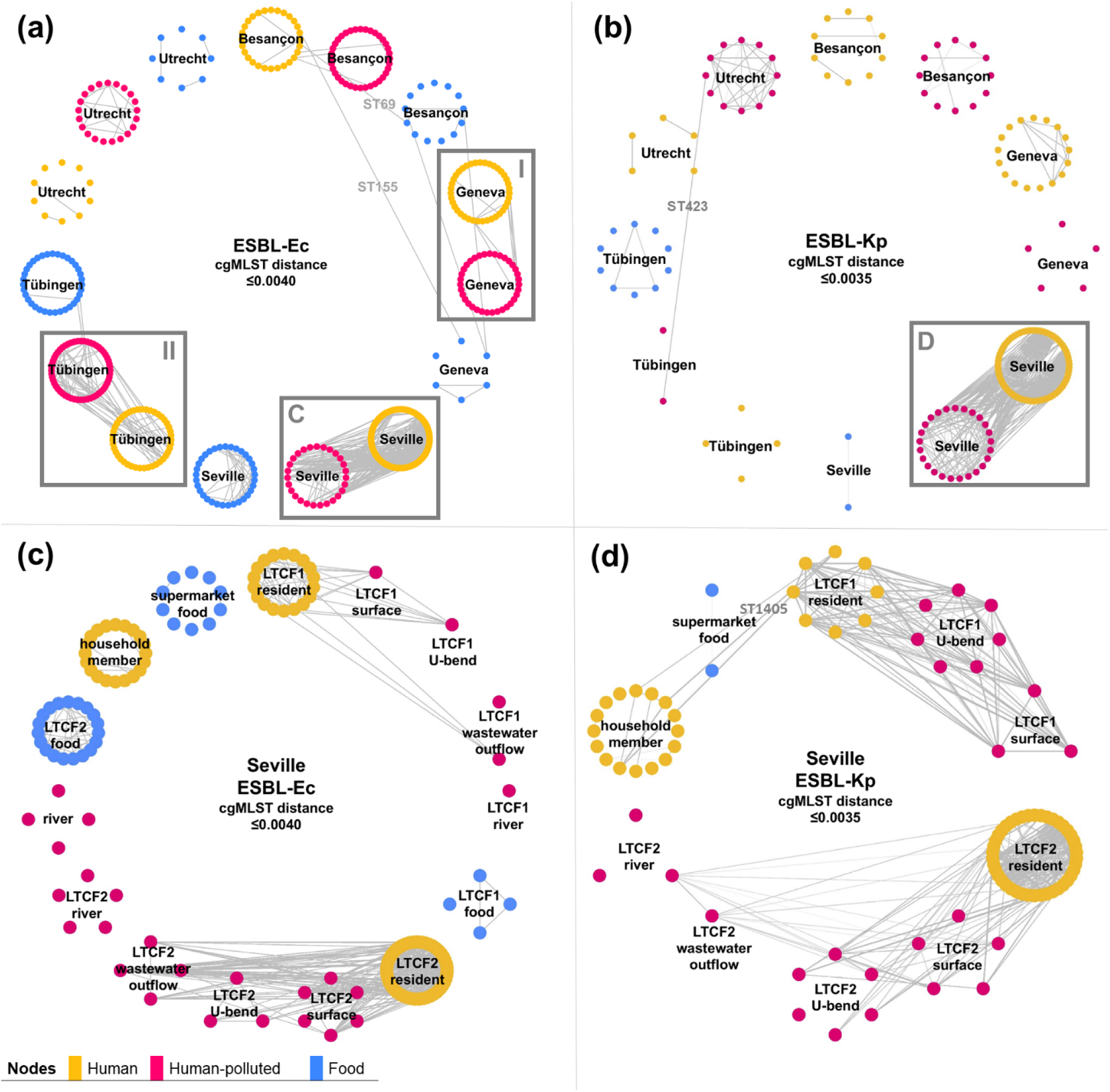
Visualization of clonally related isolate pairs. Nodes represent isolates and are grouped based on epidemiological setting, lines represent genetically similar isolate pairs. **Panel A)** ESBL-producing *E. coli* (ESBL-Ec), **Panel B)** ESBL-producing *K. pneumoniae* (including seven *K. variicola isolates*) (ESBL-Kp), **Panel C)** ESBL-Ec Seville, **Panel D)** ESBL-Kp Seville. Grey boxes I and II are displayed enlarged in S1: Fig S5, I: Panel A, II panel B.

The ML-tree of ESBL-Ec showed no evident clustering based on sample group, with the exception of the absence of food isolates within the ST131 and ST1193 phylogeny (Figure 4a, https://microreact.org/project/wSXSAfjgAH1Xr9af46Hbcb-modern-project-esbl-e-coli-n475). Both STs were retrieved in all cities, potentially indicating clonal human-to-human spread in a large geographic area. For ST131, clade A (24/138), B (5/138), and C (108/138) clustered separately, and most of these isolates carried the *bla*_CTX-M-15,_ *bla*_CTX-M-27,_ or *bla*_CTX-M-14_ *bla*_ESBL_ (Figure 4a). Clade C1 and C2 were distributed evenly with 51 and 57 isolates, respectively. Most of C1 isolates carried *bla*_CTX-M-27_ (*n*=40). For ESBL-Kp, most STs were specific to a metropolitan area, likely reflecting local clonal dissemination (Figure 4b, https://microreact.org/project/vRJHCdos8zWcv4yHMTKjak-modern-project-esbl-k-pneumoniae). However, ST405 and ST323 were observed in two countries, and ST219 (*n*=6) was observed in four countries from human, river, and chicken and turkey isolates, potentially reflecting widespread clonal dissemination. The few food-related isolates (*n*=13) were scattered throughout the tree (Figure 4b).

**Figure 4.**
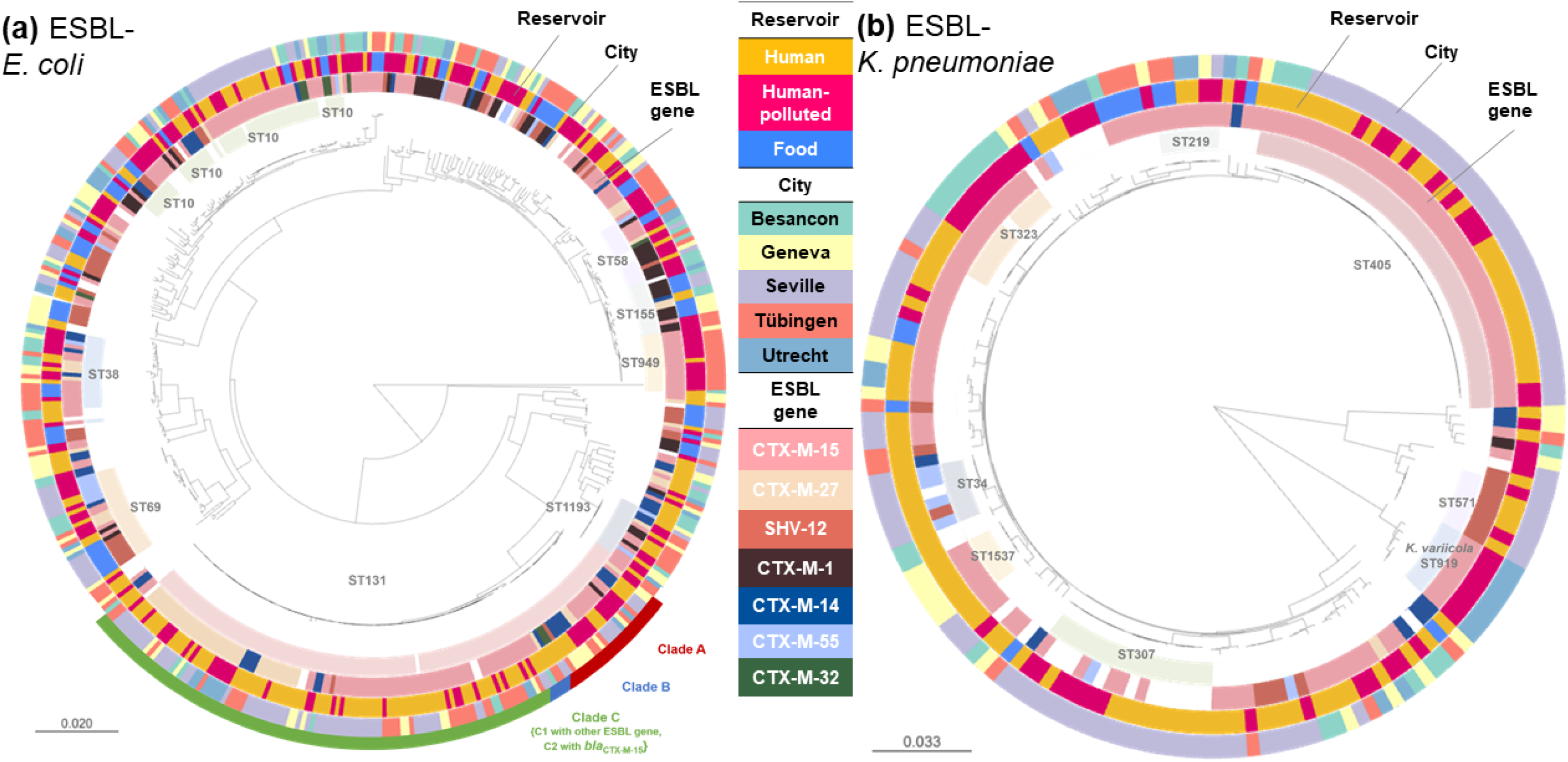
Maximum likelihood core genome phylogenies. **Panel A)** ESBL-producing *E. coli* (ESBL-Ec) (*n*=475 isolates), **Panel B)** ESBL-producing *K. pneumoniae* (ESBL-Kp) (*n=*171 isolates, including seven *K. variicola* isolates). Online access interactive trees: ESBL-Ec: https://microreact.org/project/wSXSAfjgAH1Xr9af46Hbcb-modern-project-esbl-e-coli-n475 ESBL-Kp: https://microreact.org/project/vRJHCdos8zWcv4yHMTKjak-modern-project-esbl-k-pneumoniae.

## DISCUSSION

In this multicentre prospective study, ESBL-Ec and ESBL-Kp regularly co-occurred in humans who, by the design of this study, were in close contact with healthcare. The majority of *bla*_ESBL_ types were found in both species. *bla*_ESBL_ distributions of ESBL-Ec and ESBL-Kp sampled from humans were more similar than ESBL-Ec sampled from humans and food. Non-ESBL-ARGs occurrence was highest in ESBL-Kp from any source and lowest in ESBL-Ec from food. While ESBL-Kp did more often carry ARGs, substantial overlap was observed for ARGs encoding resistance to beta-lactams, and aminoglycosides between ESBL-Ec and ESBL-Kp from humans.

A relatively, low similarity in *bla*_ESBL_ type distributions was observed between ESBL-Ec from food and humans, which was mainly explained by a lower occurrence of *bla*_CTX-M-15_, and higher occurrence of *bla*_CTX-M-1_ and *bla*_SHV-12_ in food [4]. Furthermore, ST131, the most prevalent ST in the dataset, was absent in food, and ARG proportions were lowest in food-related ESBL-Ec. Early research showed potential links between ESBL-Ec from food and humans [5]. However, more recent studies found that, at least in high income countries, ESBL-Ec and ESBL-Kp mainly spread through human-to-human transmission, while transmission of ESBL-Ec from food is less frequently observed and does probably not play an important role for ST131 the most frequently observed ST in humans. The role of food can be considered as spill-over events (potentially introducing new resistance traits into humans which may subsequently spread) [4, 6, 22].

The *bla*_CTX-M-15_ gene has been widely observed as the most frequently occurring *bla*_ESBL_ in both *E. coli* and *K. pneumoniae* from human carriage and infection [4, 6, 19]. In this study, the majority of the detected *bla*_ESBL_ types occurred in both species, with *bla*_CTX-M-15_ detected in more than half of the included isolates. Furthermore, we observed considerable similarity in ARG content encoding resistance to beta-lactams, aminoglycosides, and sulfonamides. These observations are supported by studies that statistically compared ARG content between *E. coli* and *K. pneumonia*e, demonstrating overlap in ARGs conferring resistance to several antimicrobial classes [13–17].

To our knowledge this is the first study to genetically compare ESBL-Ec and ESBL-Kp that were prospectively and longitudinally sampled from humans, the hp-environment, and food in five European metropolitan areas. Due to the sampling strategy we were able to give unique insight in the co-occurrence of these species. Furthermore, the availability of short read sequences of all isolates enabled us to compare the assessed reservoirs on several genetic levels.

Several limitations need to be considered. Firstly, the results of this study could not assess the HGT of ARGs, as short read sequencing technology is not a reliable in terms of analysing mobile genetic elements such as plasmids [23]. This requires a transmission analysis using long read sequencing, which will be performed in future MODERN studies. Secondly, the restricted sampling frame has significant limitations in accurately reflecting the ARG overlap. Since the samples were taken at defined points in time, there was an asymmetry in terms of the number of samples per reservoir as shown in S1: Figure S1. In addition, the sample size of ESBL-Kp was considerably smaller than that of ESBL-Ec due to a lower prevalence of this species, which may have hampered comparison of the two species. ESBL-Kp from food was particularly rare, prohibiting us from drawing any conclusions about this variant in relation to this reservoir. Thirdly, we used two different cgMLST thresholds, one to select unique intra-individual isolates, and one to determine clonal tranmission between humans and non-human groups. Using the same threshold would have overestimated either the number of unique intra-individual isolates, or the clonal relationships between humans and non-human groups. The optimal threshold for the sampled groups is unknown, and likely differs per situation. Fourthly, although LTCF-associated food items were sampled directly from the LCTFs, household-associated food was sampled from supermarkets, and thus, not directly related to their food intake. A higher similarity between ESBL-Ec from human carriage and food may have been observed if the food was sampled directly from the households, and on more time-points. As a consequence, this study could not reliably determine the exact relation between contamination of specific food items and ESBL carriage in the participating individuals. Lastly, the generalisability of the results to community dwellers is likely low, as we included populations closely related to healthcare. In these settings the prevalence of ESBL-Ec, and ESBL-Kp particularly, is higher than in the general community. This inherently resulted in a higher likelihood of human-to-human clonal transmission and HGT, likely leading to overestimation of the similarity of ARGs of ESBL-Ec and ESBL-Kp in humans in general.

Our data suggest the importance of public health interventions on prevention of human-to-human clonal and horizontal transmission through (i) implementation of adequate infection prevention in healthcare centers like LTCFs, and (ii) development of hygienic advice for households of colonised patients discharged from hospitals. Future research should elucidate to which extent, and under which circumstances, members of the *Enterobacterales* family share ARGs through HGT, and how this contributes to endemicity of ESBL-Ec and ESBL-Kp.

In conclusion, ESBL-Ec and ESBL-Kp regularly co-occurred in human populations with a close relationship with healthcare throughout Europe. Considerable overlap in ARG content was observed between ESBL-Ec and ESBL-Kp. Furthermore, clonal links were frequently observed between humans and the human-polluted environment, and at a lower frequency between humans and food. These findings are consistent with human-to-human transmission as an important driver of the dissemination of ARGs in humans.

## Data Availability

Raw reads are available on the European Nucleotide Archive (ENA), under the accession number PRJEB50545. Metadata with accession numbers are available in the supplementary material. Online access interactive maximum-likelihood trees: ESBL-Ec: https://microreact.org/project/wSXSAfjgAH1Xr9af46Hbcb-modern-project-esbl-e-coli-n475, ESBL-Kp: https://microreact.org/project/vRJHCdos8zWcv4yHMTKjak-modern-project-esbl-k-pneumoniae.

https://www.ebi.ac.uk/ena/browser/view/PRJEB50545

https://microreact.org/project/wSXSAfjgAH1Xr9af46Hbcb-modern-project-esbl-e-coli-n475

https://microreact.org/project/vRJHCdos8zWcv4yHMTKjak-modern-project-esbl-k-pneumoniae

## ACKNOWLEDGEMENTS

We would like to thank the staff and residents from the LTCFs of Bellevaux in Besançon – France; Ferrusola and El Recreo in Seville – Spain; Luise-Wetzel-Stift and Samariterstift in Tübingen, Germany; and Altenahove in Almkerk – The Netherlands for their involvement in the study. We would like to thank Mercedes Delgado from Seville – Spain; Caroline Brossier from Geneva - Switzerland, for assistance in collecting and processing the samples. We would like to thank Judith Vlooswijk, and Heike Schmitt from Utrecht – the Netherlands; Marion Broussier, Jeanne Celotto, and Alexandre Meunier from Besançon – France; John Poté, Gesuele Renzi, Abdessalam Cherkaoui, and Siva Lingam from Geneva, Switzerland; Michael Eib from Tübingen – Germany for their help on microbiological analyses of the samples, Elisabeth Stoll and Steffen Ganß from Tübingen – Germany for collecting the samples. We thank the Institut Pasteur teams for the curation and maintenance of BIGSdb-Pasteur databases at http://bigsdb.pasteur.fr/. We thank the Enterobase team for the curation and maintenance of Enterobase *Escherichia coli* MLST database at https://enterobase.warwick.ac.uk.

## TRANSPARENCY DECLARATIONS

DH, JAJWK, SH, JRB, ET, IBA, and BSC conceptualised the study, and obtained funding. TDV, JG, MER, DM, DH, ET, SH, ACF, SP, and JAJWK wrote the study protocols, for epidemiologic data collection, microbiologic procedures, and whole genome sequencing procedures. DM, TDV, NC, SG, MER, and ES collected the samples. DM, SP, JS, ACF, JG, and DH performed or supervised the microbiological analyses in their local centres. JG, SP, JS, ACF, TDV, JAJWK performed and supervised sequencing. TDV, JG, DM, MER, and JS verified the underlying data. TDV and JG performed genetic and statistical analyses. DH, JAJWK, SH, JRB, SG, and ET supervised the study in their local institution as principal investigators. TDV, JG, SP, ACF, and JAJWK drafted the manuscript and all authors reviewed and edited the manuscript.

All authors declare no competing interest. All authors had full access to all the data in the study and had final responsibility for the decision to submit for publication.

## REFERENCES

1. World Health Organization. Thirteenth general programme of work. 2019. https://apps.who.int/iris/bitstream/handle/10665/324775/WHO-PRP-18.1-eng.pdf.

2. Cassini, A, Högberg LD, Plachouras D et al. Attributable deaths and disability-adjusted life-years caused by infections with antibiotic-resistant bacteria in the EU and the European Economic Area in 2015: a population-level modelling analysis. Lancet Infect Dis 2019; 1: 56–66.

3. Willems RPJ, van Dijk K, Vehreschild MJGT et al. Incidence of infection with multidrug-resistant Gram-negative bacteria and vancomycin-resistant enterococci in carriers: a systematic review and meta-regression analysis. Lancet Infect Dis 2023; 6: 719–731.

4. Dorado-García A, Smid JH, van Pelt W et al. Molecular relatedness of ESBL/AmpC-producing *Escherichia coli* from humans, animals, food and the environment: a pooled analysis. J Antimicrob Chemother 2018; 73: 339–47.

5. Kluytmans JAJW, Overdevest ITMA, Willemsen I et al. Extended-spectrum β-lactamase-producing *Escherichia coli* from retail chicken meat and humans: comparison of strains, plasmids, resistance genes, and virulence factors. Clin Infect Dis 2013; 56: 478–87.

6. Ludden C, Moradigaravand D, Jamrozy D et al. A one health study of the genetic relatedness of *Klebsiella pneumoniae* and their mobile elements in the east of England. Clin Infect Dis 2020; 2: 219–26.

7. Martak D, Guther J, Verschuuren TD et al. Populations of extended-spectrum β-lactamase-producing *Escherichia coli* and *Klebsiella pneumoniae* are different in human-polluted environment and food items: a multicentre European study. Clin Microbiol Infect 2021; 21: 00414–6.

8. Hendrickx APA, Landman F, de Haan A et al. blaOXA-48-like genome architecture among carbapenemase-producing *Escherichia coli* and *Klebsiella pneumoniae* in the Netherlands. Microb Genom 2021; 5: 000512.

9. Nang SC, Li J, Velkov T. The rise and spread of mcr plasmid-mediated polymyxin resistance. Crit Rev Microbiol 2019; 2: 131–61.

10. Abdel-Rahim MH, El-Badawy O, Hadiya S et al. Patterns of fluoroquinolone resistance in Enterobacteriaceae isolated from the Assiut university hospitals, Egypt: A comparative study. Microb Drug Resist 2019; 4: 509–19.

11. Li P, Liu D, Zhang X et al. Characterization of plasmid-mediated quinolone resistance in gram-negative bacterial strains from animals and humans in China. Microb Drug Resist 2019; 7: 1050–6.

12. Mitra S, Mukherjee S, Naha S et al. Evaluation of co-transfer of plasmid-mediated fluoroquinolone resistance genes and bla NDM gene in Enterobacteriaceae causing neonatal septicaemia. Antimicrob Resist Infect Control 2019; 8: 46.

13. Zhang Y, Yang J, Ye L et al. Characterization of clinical multidrug-resistant *Escherichia coli* and *Klebsiella pneumoniae* isolates, 2007-2009, China. Microb Drug Resist 2012; 5: 465–70.

14. Zhou Z, Zhang K, Chen W et al. Epidemiological characteristics of carbapenem-resistant Enterobacteriaceae collected from 17 hospitals in Nanjing district of China. Antimicrob Resist Infect Control 2020; 1: 15.

15. Riccio ME, Verschuuren T, Conzelmann N et al. Household acquisition and transmission of extended-spectrum β-lactamase (ESBL)-producing Enterobacteriaceae after hospital discharge of ESBL-positive index patients. Clin Microbiol Infect 2021; 20: 30784–9.

16. Zankari E, Hasman H, Cosentino S et al. Identification of acquired antimicrobial resistance genes. J Antimicrob Chemother 2012; 11: 2640–4.

17. Breakpoint tables for interpretation of MICs and zone diameters, version 13, 2022, The European Committee on Antimicrobial Susceptibility Testing, https://www.eucast.org/fileadmin/src/media/PDFs/EUCAST_files/Breakpoint_tables/v_13.0_Breakpoint_Tables.xlsx.

18. Bortolaia V, Kaas RS, Ruppe E et al. ResFinder 4.0 for predictions of phenotypes from genotypes. J Antimicrob Chemother 2020; 12: 3491–3500.

19. Verschuuren TD, van Hout D, Arredondo-Alonso S et al. Comparative genomics of ESBL-producing Escherichia coli (ESBL-Ec) reveals a similar distribution of the 10 most prevalent ESBL-Ec clones and ESBL genes among human community faecal and extra-intestinal infection isolates in the Netherlands (2014–17). J Antimicrob Chem 2021; 4: 901–8.

20. Larsen MV, Cosentino S, Rasmussen S et al. Multilocus sequence typing of total-genome-sequenced bacteria. J Clin Microbiol 2012; 4: 1355–61.

21. Wolda H. Similarity indices, sample size and diversity. Oecologia 1981; 50: 296–302.

22. Perestrelo S, Carreira GC, Valentin L et al. Comparison of approaches for source attribution of ESBL-producing Escherichia coli in Germany. PLoS One 2022; 7: e0271317.

23. Arredondo-Alonso S, Willems RJ, van Schaik W, Schürch AC. On the (im)possibility of reconstructing plasmids from whole-genome short-read sequencing data. Microb Genom 2017; 10: e000128.

