## Supplementary material for "Extended-spectrum beta-lactamase-producing *Escherichia coli* and *Klebsiella pneumoniae* from human carriage, the human-polluted environment, and food: molecular epidemiology of two prospective cohorts in five European metropolitan areas": S1

### Supplementary appendix

### METHODS

#### Appendix 1: Sample composition

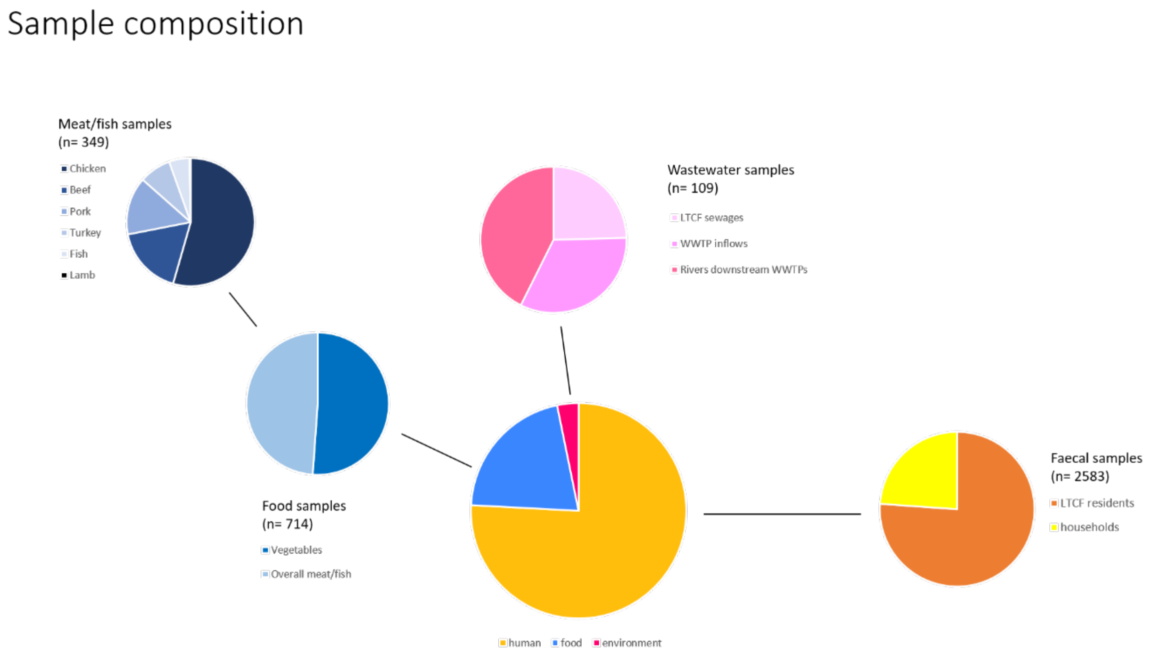

**Figure S1 (a). Composition of all collected samples**

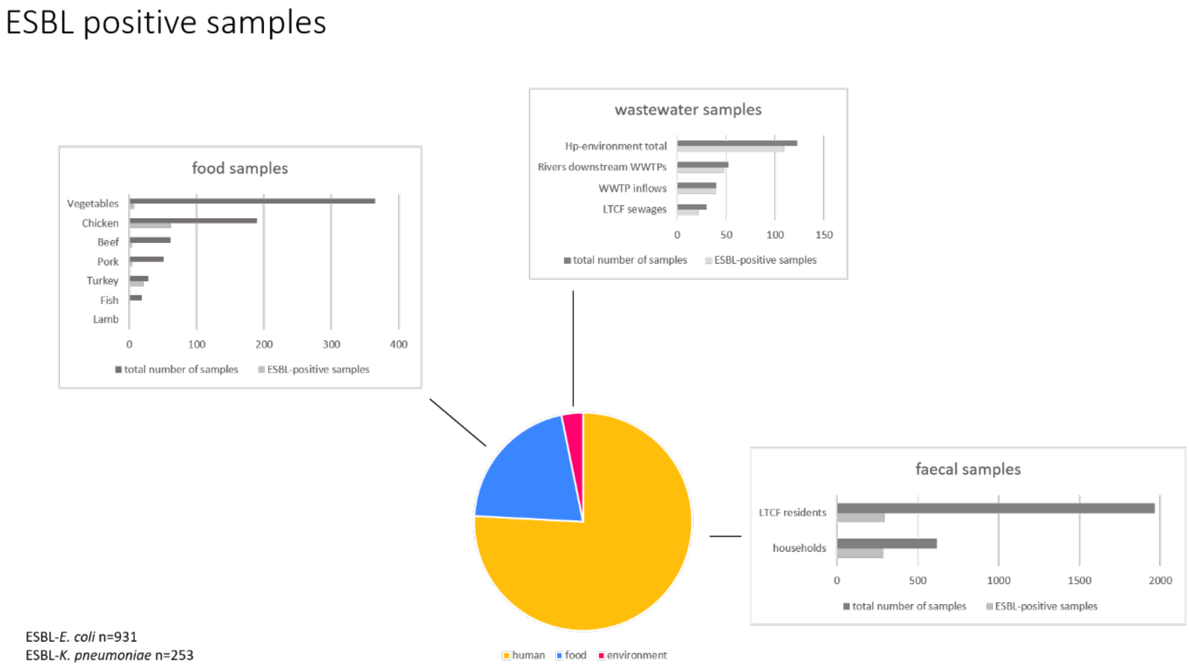

**Figure S1 (b). Composition of ESBL positive samples**

**Appendix 2: Microbiological methods**

Stool samples, rectal swabs, and environmental surface samples were streaked directly on ChromID ESBL agar (bioMérieux, Marcy l'Etoile, France). Additionally, MacConkey broth with vancomycin 64 µg/mL and 32 µg/mL cefuroxime, incubated for 24 h at 35°C. Ten µL of the broth was subsequently streaked on ChromID ESBL agar and further incubated for 48 h at 35°C.

Samples from wastewater treatment plants were diluted 1:10 in sterile water, 10 µL and 100 µL were streaked on ChromID agar and incubated overnight at 35°C. One hundred millilitres of river and LTCF sewage samples were filtered on a 0.45-µm filter deposited on ChromID agar and incubated overnight at 35°C.

Food samples, 25 g of meat or vegetables were incubated overnight at 35°C in 250 mL of tryptic soy broth supplemented with 8 mg/L vancomycin and 0.25 mg/L cefotaxime. Then, 100 µL was streaked on ChromID agar and incubated overnight at 35°C.

Per plate, each unique colony morphology suspected to be *E. coli* or *K. pneumoniae* was identified using Matrix Assisted Laser Desorption Ionization - Time of Flight Mass Spectrometry (MALDI-TOF MS). ESBL production was confirmed by double disk synergy tests (DDST20 and DDST30) and by the determination of the beta-lactamase inhibition profile (ESBL + AmpC Screen ID Kit, Rosco Diagnostica, Taastrup, Denmark). Based on distinct colony morphology, each centre stored isolates at -80 degrees Celsius 1 to 4 isolates per sample in beadcontaining cryotubes (Microbank, PRO-LAB Diagnostics, ON, Canada) until further analysis to prevent plasmid loss.

**Appendix 3: Quality assessment of sequences and included isolates**

Quality of assembled sequences was assessed by inspecting the number of contigs per assembly, the total genome size, and the GC content. In this study only unique isolates were included, defined as the first isolate per participant or non-human sample of one of the two species. A second isolate was included if the cgMLST pairwise genetic distance was >0.0105 (~25 alleles) for ESBL-Ec, or >0.0035 (~10 alleles) for ESBL-Kp [1]. Distance matrices were generated (Ridom Seqsphere v5.0), using the Enterobase scheme (2,513 target genes) for ESBL-Ec, and the *K. pneumoniae/variicola/quasipneumoniae* sensu lato scheme (2,358 target genes) for EBSL-Kp [2, 3]. Pairwise distance was expressed as the proportion of allele differences ( $\frac{\text{\# allele differences}}{\text{\# good target gene shared}}$ ).

**Appendix 4: *In silico* molecular typing**

Sequence types (STs) were identified using tseemann/mlst (v2.16.2) (https://github.com/tseemann/mlst) [4]. Sub-clades were assigned to ST131 (ESBL-Ec) isolates: clade A (*fimH*41), clade B (*fimH*22), clade C (*fimH*30 – non-FQR), clade C1 (*fimH*30 - FQR-non *bla*CTX-M-15), and lastly clade C2 (*fimH*30 - FQR-*bla*CTX-M-15) [5]. Observed other *fimH* types were assigned to a clade according to their position in the phylogenetic tree [6].

Antimicrobial resistance genes (ARGs) were identified with Resfinder (v3.2) using abricate (v0.8.10), with a length of ≤80% and an identity of ≤95% [7]. Of the in total 4,344 detected ARGs, 68% (*n*=2,945) had an identical match in length and identity. 93% (*n*=4,037) of the detected ARGs had an identical match in length with the target ARG (Figure below, left), while 70% (*n*=3,025) of the detected ARGs had an identical match in identity with the target ARG (Figure below, right). Chromosomal combined *gyrA/parC* mutations conferring fluoroquinolone resistance (FQR) were determined with PointFinder (v4.1) [8]. *FimH* types were assigned using FimTyper (v1.0) [9].

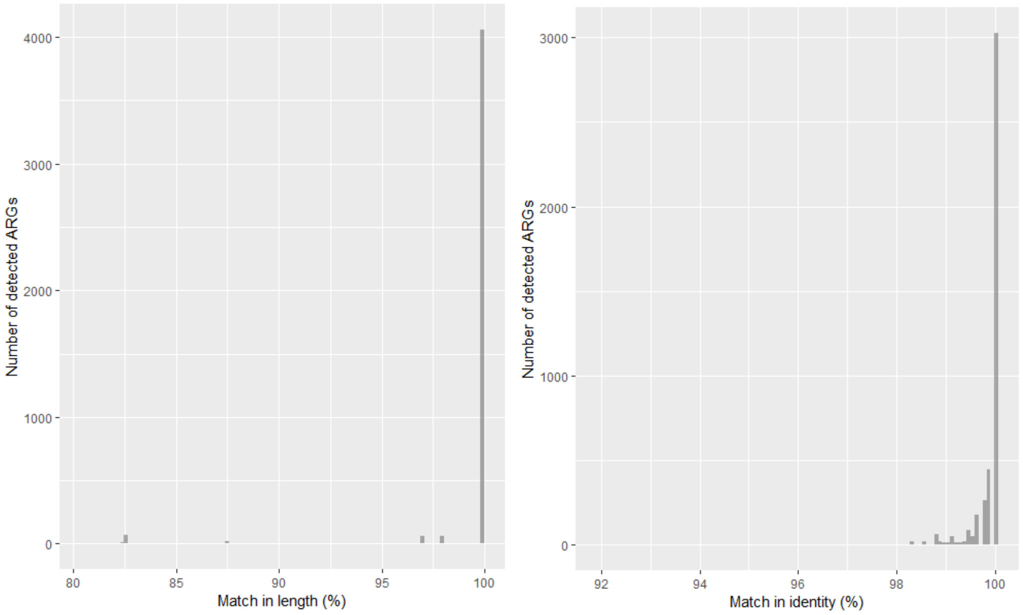

**Figure S2. Histogram of match in length (left), and match in identity (right) of detected antimicrobial resistance genes with ResFinder (v3.2)**

**Appendix 5: Genotype to phenotype translation table of ARGs**

This table was based on the following genotype-to-phenotype translation table of ResFinder (https://bitbucket.org/genomicepidemiology/resfinder_db/src/master/phenotypes.txt), and EUCAST Clinical Breakpoint Tables (v12.0) to further subgroup the ‘β-lactam’ class.

| ARG class | Antimicrobial | Observed ARG-types[a] |
|---|---|---|
| beta-lactamases (penicillins (small- or broad-spectrum) | amoxicillin, piperacillin, amoxicillin-clavulanic acid, piperacillin-tazobactam | *bla*$_{\text{CARB-2}}$, *bla*$_{\text{DHA-1}}$, *bla*$_{\text{LEN24}}$, *bla*$_{\text{OKP-A-12}}$, *bla*$_{\text{OKP-A-3}}$, *bla*$_{\text{OKP-B-1}}$, *bla*$_{\text{OKP-B-15}}$, *bla*$_{\text{OKP-B-3}}$, *bla*$_{\text{OKP-B-9}}$, *bla*$_{\text{OXA-1}}$[b], *bla*$_{\text{OXA-2}}$, *bla*$_{\text{SCO-1}}$, *bla*$_{\text{SHV-1}}$, *bla*$_{\text{SHV-11}}$, *bla*$_{\text{SHV-110}}$, *bla*$_{\text{SHV-145}}$, *bla*$_{\text{SHV-172}}$, *bla*$_{\text{SHV-187}}$, *bla*$_{\text{SHV-27}}$, *bla*$_{\text{SHV-62}}$, *bla*$_{\text{SHV-75}}$, *bla*$_{\text{SHV-76}}$, *bla*$_{\text{TEM-1}}$, *bla*$_{\text{TEM-122}}$, *bla*$_{\text{TEM-2}}$, *bla*$_{\text{TEM-210}}$ *bla*$_{\text{TEM-33}}$ |
| ESBL (penicillin + cephalosporin) | amoxicillin, aztreonam, cefepime, cefotaxime, ceftazidime, ceftriaxone, piperacillin | *bla*$_{\text{CTX-M-1}}$, *bla*$_{\text{CTX-M-14}}$, *bla*$_{\text{CTX-M-15}}$, *bla*$_{\text{CTX-M-27}}$, *bla*$_{\text{CTX-M-209}}$, *bla*$_{\text{CTX-M-3}}$, *bla*$_{\text{CTX-M-32}}$, *bla*$_{\text{CTX-M-55}}$, *bla*$_{\text{CTX-M-65}}$, *bla*$_{\text{CTX-M-8}}$, *bla*$_{\text{CTX-M-9}}$, *bla*$_{\text{SHV-106}}$, *bla*$_{\text{SHV-12}}$, *bla*$_{\text{SHV-129}}$, *bla*$_{\text{SHV-13}}$, *bla*$_{\text{SHV-2}}$, *bla*$_{\text{SHV-5}}$, *bla*$_{\text{TEM-169}}$, *bla*$_{\text{TEM-106}}$, *bla*$_{\text{TEM-52}}$ |

| | | |
|---|---|---|
| carbapenemase (penicillin + carbapenem) | amoxicillin, amoxicillin-clavulanic acid, imipenem, meropenem, piperacillin, piperacillin-tazobactam | *bla*$_{\text{KPC-2}}$[c], *bla*$_{\text{OXA-181}}$[d], *bla*$_{\text{OXA-48}}$ |
| fluoroquinolones | ciprofloxacin | *aac(6')-Ib-cr, oqxA, oqxB, qepA4, qnrA1, qnrB1, qnrB19, qnrB4, qnrB52, qnrS1, qnrS2* |
| aminoglycosides | amikacin, gentamicin, tobramycin, streptomycin, neomycin, hygromycin | *aac(3)-IIa, aac(3)-IId, aac(3)-Iva, aac(3)-Via, aac(6')-Ib-cr, aadA1, aadA16, aadA2, aadA4, aadA5, ant(2'')-Ia, ant(3'')-Ia, aph(3')-Ia, aph(3'')-Ib, aph(3')-Id, aph(4)-Ia, aph(6)-Id* |
| fosfomycin | fosfomycin | *fosA, fosA3, fosA5, fosA6* |
| trimethoprim | trimethoprim | *dfrA1, dfrA12, dfrA14, dfrA15, dfrA16, dfrA17, dfrA27, dfrA5, dfrA7, dfrA8* |
| sulfonamides | sulfonamides | *sul1, sul2, sul3* |
| colistin | colistin | *mcr-1* |

[a]ARG does not need to confer resistance to all antimicrobials listed in their respective antimicrobial group
[b]also predicted to confer resistance to cefepime
[c]also predicted to confer resistance to cefepime, cefotaxime, cefoxitin, ceftazidime, and ertapenem
[d]also predicted to confer resistance to cefepime, and ertapenem

**Appendix 6: maximum likelihood phylogenetic trees**

Maximum likelihood (ML) trees were created for ESBL-Ec and ESBL-Kp with Spine v0.1.2 [10]. Prophage sequences were identified using phaster.ca and removed [11]. Single nucleotide polymorphism (SNP) calling was performed with a customized python pipeline, which includes GATK v.3.2-2 and SAMtools v.0.1.19 [12, 13]. The ML-trees were calculated with RAxML v.8.2.6 with a general time-reversible (GTR) substitution model and a gamma distribution of rates undergoing 1,000 bootstraps [14]. ML-trees were visualised using microreact (v.218) [15].

### RESULTS

**Table S1. Description of participants carrying multiple ESBL-*E. coli* ST131 strains over time**. Definition unique clone: Isolates within the same participant with a cgMLST distance greater than 0.0105[1]

| City | Participant | Clade | $bla_{ESBL}$ | cgMLST dist[a] |
|---|---|---|---|---|
| Seville | LTCF2 resident | H30R (clade C2) | $bla_{CTX-M-15}$ | - |
| Seville | LTCF2 resident | H30R (clade C1) | $bla_{CTX-M-27}$ | 0.0173 |
| Seville | LTCF2 resident | H30R (clade C1) | $bla_{CTX-M-27}$ | - |
| Seville | LTCF2 resident | H30R (clade C2) | $bla_{CTX-M-15}$ | 0.0205 |
| Seville | LTCF2 resident | H41 (clade A) | $bla_{CTX-M-14}$ | - |
| Seville | LTCF2 resident | H30R (clade C1) | $bla_{CTX-M-14}$ | 0.2856 |
| Seville | LTCF2 resident | H41 (clade A) | $bla_{CTX-M-14}$ | - |
| Seville | LTCF2 resident | | $bla_{CTX-M-27}$ | 0.2833 (versus 0.3187.1) |
| Seville | LTCF2 resident | H30R (clade C2) | $bla_{CTX-M-15}$ | 0.2894 (versus 0.3187.1)<br>0.0209 (versus 0.3187.2) |
| Seville | LTCF1 resident | H30R (clade C1) | $bla_{CTX-M-27}$ | - |
| Seville | LTCF1 resident | H41 (clade A) | $bla_{CTX-M-15}$ | 0.2729 |
| Seville | LTCF1 resident | H30R (clade C1) | $bla_{CTX-M-27}$ | - |
| Seville | LTCF1 resident | H41 (clade A) | $bla_{CTX-M-15}$ | 0.2737 |
| Tübingen | Index household | H30R (clade C2) | $bla_{CTX-M-15}$<br>$bla_{CTX-M-27}$ | - |
| Tübingen | Index household | H54 (clade B) | $bla_{CTX-M-27}$ | 0.0665 (versus 21656.2) |
| Tübingen | Index household | H54 (clade B) | $bla_{CTX-M-14}$ | 0.0667 (versus 21656.2)<br>0.0226 (versus 21672.2) |
| Tübingen | LTCF1 resident | H30R (clade C1) | $bla_{CTX-M-14}$ | - |
| Tübingen | LTCF1 resident | H30R (clade C1) | $bla_{CTX-M-14}$ | 0.0228 (versus 1.1539.1) |
| Tübingen | LTCF1 resident | H30R (clade C1) | $bla_{CTX-M-14}$ | 0.0220 (versus 1.1539.1)<br>0.0136 (versus 1.1749.1) |

[a]Pairwise cgMLST distance, calculated with Enterobase scheme available in Seqsphere (v5.0)

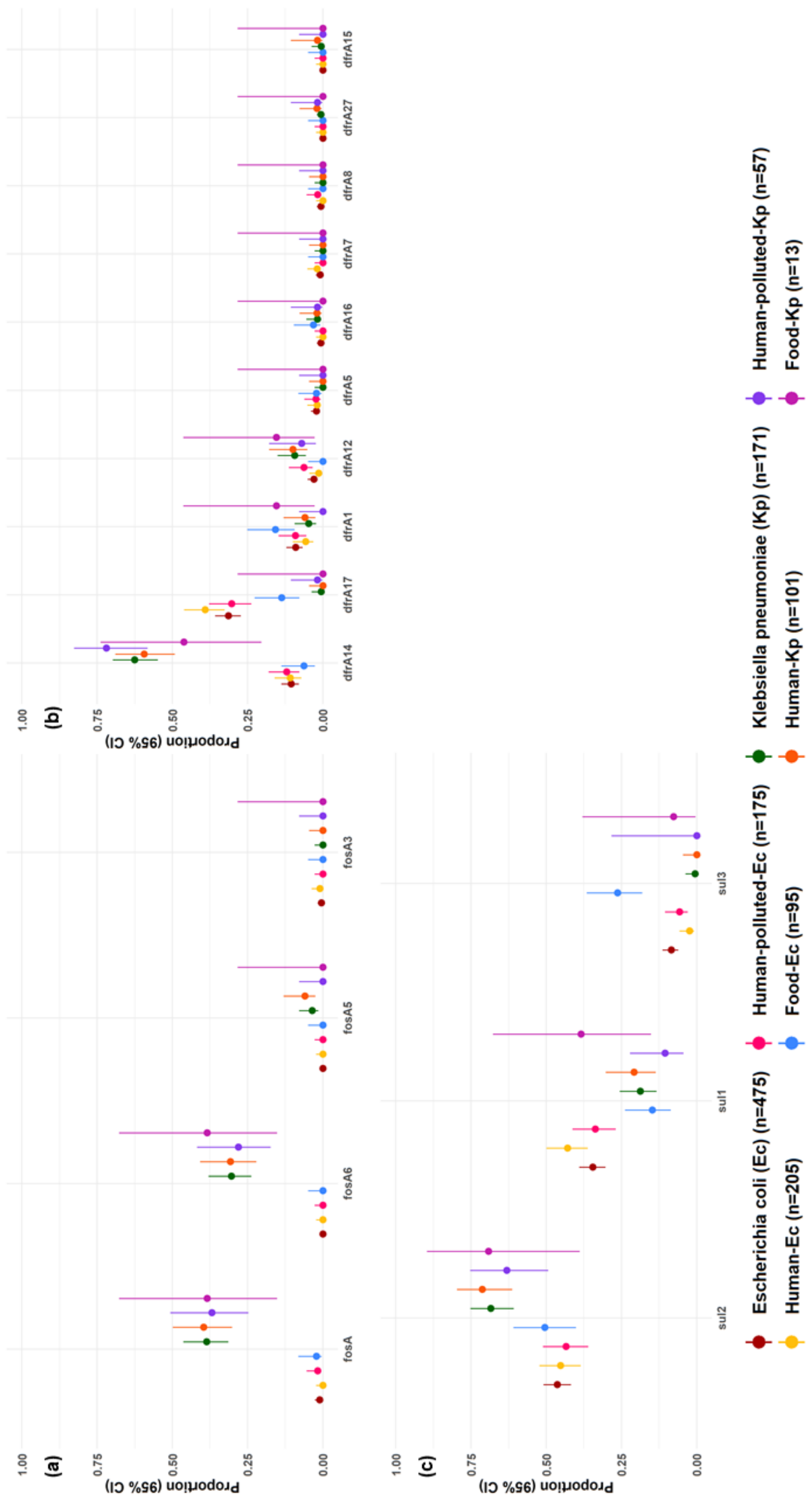

**Figure S3. Proportions (95% CI) of the ten most frequent antimicrobial resistance gene types per isolate.**

**Panel A)** fosfomycin, **Panel B)** trimethoprim, **Panel C)** sulfanomides.

| (a) | EC H | EC HP | EC F | KP H | KP HP | KP F |
|---|---|---|---|---|---|---|
| EC H | - | 0.94 .9-1. | 0.94 .9-1. | 0.24 .1-.3 | 0.35 .2-.4 | 0.23 .0-.5 |
| | EC HP | - | 0.94 .9-1. | 0.26 .1-.4 | 0.37 .2-.5 | 0.25 .0-.5 |
| | | EC F | - | 0.26 .1-.4 | 0.37 .2-.5 | 0.25 .0-.5 |
| | | | KP H | - | 0.83 .7-.9 | 0.76 .6-.9 |
| | | | | KP HP | - | 0.76 .5-.8 |

| (b) | EC H | EC HP | EC F | KP H | KP HP | KP F |
|---|---|---|---|---|---|---|
| EC H | - | 0.85 .8-.9 | 0.67 .5-.8 | 0.36 .3-.5 | 0.31 .2-.4 | 0.38 .1-.6 |
| | EC HP | - | 0.72 .6-.8 | 0.42 .3-.5 | 0.35 .2-.5 | 0.45 .2-.7 |
| | | EC F | - | 0.34 .2-.5 | 0.27 .1-.4 | 0.40 .1-.7 |
| | | | KP H | - | 0.81 .7-.9 | 0.72 .5-.9 |
| | | | | KP HP | - | 0.65 .4-.9 |

| (c) | EC H | EC HP | EC F | KP H | KP HP | KP F |
|---|---|---|---|---|---|---|
| EC H | - | 0.88 .8-.9 | 0.72 .6-.8 | 0.75 .6-.9 | 0.71 .6-.8 | 0.75 .5-.9 |
| | EC HP | - | 0.74 .6-.8 | 0.73 .6-.8 | 0.74 .6-.9 | 0.71 .5-.9 |
| | | EC F | - | 0.73 .6-.8 | 0.73 .6-.8 | 0.68 .5-.9 |
| | | | KP H | - | 0.84 .7-.9 | 0.76 .6-.9 |
| | | | | KP HP | - | 0.68 .5-.9 |

**Legend**

| Species | |
|---|---|
| EC | ESBL-producing *E. coli* |
| KP | ESBL-producing *K. pneumoniae* |
| **Reservoir** | |
| H | human |
| HP | human-polluted environment |
| F | food |
| **Acquired resistance gene class** | |
| (a) | fosfomycin |
| (b) | trimethoprim |
| (c) | sulfonamides |

| PSI |
|---|
| 0 |
| 0.1 |
| 0.2 |
| 0.3 |
| 0.4 |
| 0.5 |
| 0.6 |
| 0.7 |
| 0.8 |
| 0.9 |
| 1 |

**Figure S4. Czekanowski's proportional similarity index (PSI) (95% confidence interval) of detected antimicrobial resistance gene (ARG) types. Panel A)** fosfomycin, **Panel B)** trimethoprim, **Panel C)** sulfonamides. The PSI is calculated with the following formula: $1 - 0.5 \sum k\, |p(reservoir[n])k - q(reservoir[nx])k|$, where $p$ corresponded to the relative frequency of gene type $k$ (e.g. *bla*$_{\text{CTX-M-15}}$) in reservoir $n$ (e.g. ESBL-*E. coli* from humans), and $q$ corresponded to the relative frequency of the same gene type in reservoir $nx$ (e.g. ESBL-*K. pneumoniae* from humans). The numerator of the relative frequency was the count of each ARG type. The denominator of the relative frequency was the total number genes of the corresponding ARG class. The PSI is a proportion, with 0 interpreted as no overlap, and 1 as perfect overlap in ARG type distributions between two reservoirs. 95% confidence intervals were calculated with 5,000 bootstrap iterations.

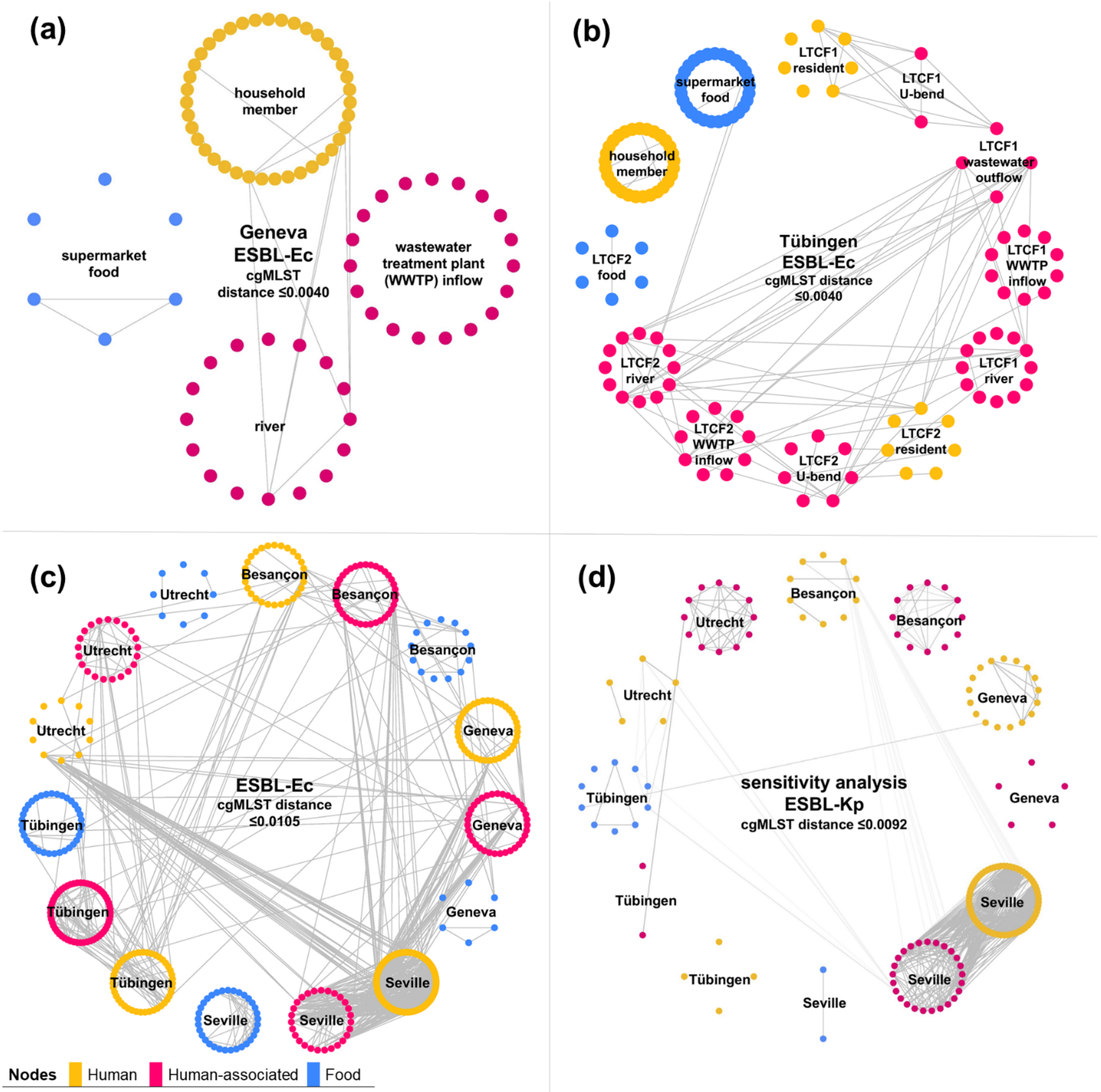

**Figure S5. Visualization of clonally related isolate pairs of ESBL-*E. coli* and ESBL-*K. pneumoniae*.** Nodes represent isolates and are grouped based on epidemiological setting, lines represent genetically similar isolate pairs. **A)** ESBL-producing *E. coli* (ESBL-Ec) Geneva, **B)** ESBL-Ec Tübingen. **C)** Sensitivity analysis with increased cgMLST distance results in connections between most reservoirs and cities for ESBL-Ec, **D)** Sensitivity analysis with increased cgMLST distance results in additional connections between most reservoirs and cities for ESBL-Kp.

#### References

1. Kluytmans-van den Bergh MFQ, Rossen JWA, Bruijning-Verhagen PCJ *et al.* Whole-genome multilocus sequence typing of extended-spectrum-B-lactamase-producing Enterobacteriaceae. *J Clin Microbiol* 2016; **12**: 2919-27.
2. Miro E, Rossen JWA, Chlebowicz MA *et al.* Core/whole genome multilocus sequence typing and core genome SNP-based typing of OXA-48-producing *Klebsiella pneumoniae* clinical isolates from Spain. *Front Microbiol* 2020; **10**: 2961.
3. Zhou Z, Alikhan NF, Mohamed K *et al*. The EnteroBase user's guide, with case studies on *Salmonella* transmissions, *Yersinia pestis* phylogeny, and *Escherichia* core genomic diversity. *Genome Res* 2020; **30**: 138-52.
4. Larsen MV, Cosentino S, Rasmussen S *et al.* Multilocus sequence typing of total-genome-sequenced bacteria. *J Clin Microbiol* 2012; **50**: 1355–61.
5. Banerjee R, Johnson JR. A new clone sweeps clean: The enigmatic emergence of Escherichia coli sequence type 131. *Antimicrob Agents Chemother* 2014; **58**: 4997–5004.
6. Verschuuren TD, van Hout D, Arredondo-Alonso S *et al*. Comparative genomics of ESBL-producing Escherichia coli (ESBL-Ec) reveals a similar distribution of the 10 most prevalent ESBL-Ec clones and ESBL genes among human community faecal and extra-intestinal infection isolates in the Netherlands (2014–17). *J Antimicrob Chem* 2021; **4**: 901–8.
7. Zankari E, Hasman H, Cosentino S *et al*. Identification of acquired antimicrobial resistance genes. J Antimicrob Chemother 2012; **67**: 2640–44.
8. Zankari E, Allesøe R, Joensen KG *et al.* PointFinder: a novel web tool for WGS-based detection of antimicrobial resistance associated with chromosomal point mutations in bacterial pathogens. *J Antimicrob Chemother* 2017; **10**: 2764-8.
9. Roer L, Tchesnokova V, Allesoe R *et al*. Development of a web tool for *Escherichia coli* subtyping based on *fimH* alleles. *J Clin Microbiol* 2017; **8**: 2538-43.
10. Ozer EA, Allen JP, Hauser AR. Characterisation of the core and accessory genomes of *Pseudomonas aeruginosa* using bioinformatic tools Spine and AGEnt. *BMC Genomics* 2014; **15**: 737.
11. Arndt D, Grant JR, Marcu A *et al.* PHASTER: a better, faster version of the PHAST phage search tool. *Nucleic Acids Res* 2016; **44**: W16-21.
12. Li H, Handsaker B, Wysoker A *et al*. The sequence alignment/map format and SAMtools. *Bioinformatics* 2009; **25**: 2078–9.
13. McKenna A, Hanna M, Banks E *et al*. The genome analysis toolkit: a mapreduce framework for analyzing next-generation DNA sequencing data. *Genome Res* 2010; **20**: 1297–303.
14. Stamatakis A. RAxML version 8: a tool for phylogenetic analysis and post-analysis of large phylogenies. *Bioinformatics* 2014; **30**: 1312–3.
15. Argimón S, Abudahab K, Goater RJE *et al*. Microreact : visualizing and sharing data for genomic epidemiology and phylogeography. *Microb Genom* 2016; **30**: 1-11.
